# Ultra-sensitive molecular residual disease detection through whole genome sequencing with single-read error correction

**DOI:** 10.1101/2024.01.13.24301070

**Authors:** Xinxing Li, Tao Liu, Antonella Bacchiocchi, Mengxing Li, Wen Cheng, Tobias Wittkop, Fernando Mendez, Yingyu Wang, Paul Tang, Qianqian Yao, Marcus W. Bosenberg, Mario Sznol, Qin Yan, Malek Faham, Li Weng, Ruth Halaban, Hai Jin, Zhiqian Hu

**Author notes:** Corresponding authors: Ruth Halaban, Department of Dermatology, Yale University School of Medicine, New Haven, CT, USA; Yale Cancer Center, Yale School of Medicine, New Haven, CT, USA,. Zhiqian Hu, Department of Gastrointestinal Surgery, Tongji Hospital Medical College of Tongji University, No. 389, Xincun Road, Putuo District, Shanghai 200065, P.R. China; Department of General Surgery, Changzheng Hospital Naval Medical University, Shanghai, P.R. China,. Hai Jin, Department of Thoracic Surgery, Shanghai Changhai Hospital, No. 168, Changhai Road, Yangpu District, Shanghai 200433, P.R. China,. These authors contributed equally to this work (Xinxing Li, Tao Liu, Antonella Bacchiocchi, Mengxing Li).

## Abstract

While whole genome sequencing (WGS) of cell-free DNA (cfDNA) holds enormous promise for molecular residual disease (MRD) detection, its performance is limited by WGS error rate. Here we introduce AccuScan, an efficient cfDNA WGS technology that enables genome-wide error correction at single read level, achieving an error rate of 4.2×10^-7^, which is about two orders of magnitude lower than a read-centric de-noising method. When applied to MRD detection, AccuScan demonstrated analytical sensitivity down to 10^-6^ circulating tumor allele fraction at 99% sample level specificity. In colorectal cancer, AccuScan showed 90% landmark sensitivity for predicting relapse. It also showed robust MRD performance with esophageal cancer using samples collected as early as 1 week after surgery, and predictive value for immunotherapy monitoring with melanoma patients. Overall, AccuScan provides a highly accurate WGS solution for MRD, empowering circulating tumor DNA detection at parts per million range without high sample input nor personalized reagents.

**One Sentence Summary:** AccuScan showed remarkable ultra-low limit of detection with a short turnaround time, low sample requirement and a simple workflow for MRD detection.

## INTRODUCTION

Molecular residual disease (MRD) refers to the small amount of cancer cells persisting after treatment (*1*). Timely and sensitive measurement of MRD is critical for recurrence risk assessment and treatment decisions (*2*). Circulating tumor DNA (ctDNA) has emerged as a promising real-time biomarker for MRD detection and monitoring. Studies have shown that the levels of cancer-specific somatic mutations in ctDNA correlate with tumor stage, burden, and response to therapy across tumor types (*3, 4*). Compared to other blood-based cancer biomarkers, such as circulating tumor cells and cancer antigens, ctDNA provides a more sensitive and specific measure of MRD (*5, 6*).

There are currently two main strategies for ctDNA-based MRD detection: 1) the tumor-naïve approach, which tests MRD samples for changes known to be enriched in cancers (*7*), such as methylation changes and common somatic mutations; 2) the tumor-informed approach, which requires a tumor sample to identify patient specific variants and then tests MRD samples for those variants (*8, 9*).

The tumor-naïve approach is logistically simple, without the need to acquire and sequence a tumor sample and uses a universal panel to test plasma samples for a cancer signal (*7, 10, 11*). While these tests offer operational convenience, they tend to have moderate sensitivity. With a methylation-based cancer detection test, Jamshidi et al claimed a 50% analytical sensitivity to a 3.1×10^-4^ circulating tumor allele fraction (cTAF) at 98% specificity (*10*). Parikh et al. had a clinical sensitivity of 55.6% to predict CRC recurrence using plasma collected at landmark time point (4 week after surgery) with a panel combining methylation and mutation signals (*7*).

The tumor informed approach utilizes tumor-specific somatic mutations from the patient tissue for MRD analysis, which is highly specific and sensitive. Factors that impact its sensitivity include the accuracy of somatic mutation calls from the tissue and plasma samples, and the total number of cfDNA molecules interrogated, which is the product of the number of somatic variants tracked and the unique molecular depth obtained through sequencing.

Tumor informed approaches can either use bespoke or off-the-shelf MRD tests. A bespoke MRD assay is designed after tumor sequence data is available and follows a limited number of variants through ultra-deep sequencing (*12, 13*). The sequencing of a bespoke panel can be exhaustive; hence the unique molecular depth is mostly limited by the amount of available input material (*12, 14–16*). For example, Signatera, a tumor-informed NGS-based multiplex PCR assay that tracks 16 personalized markers claimed 81.3%-96.1% analytical sensitivity at limit of detection (LOD) of 10^-4^ when up to 66ng of DNA was used (*15*). Tumor-informed personalized MRD assays targeting large numbers of markers and boasting error correction using unique molecular identifiers (UMI) or duplex sequencing have shown LOD below 10^-4^. (*12, 17, 18*). PhasED-seq uses multiple somatic mutations in individual DNA fragments to lower the background noise to less than 10^-6^ and claimed LOD down to the parts per million (PPM) level given enough phased variants (*19*). While the tumor informed bespoke MRD approach can achieve very high sensitivity, the requirement of a personalized design substantially increases turnaround time (TAT) and creates considerable logistical challenges.

The tumor-informed off-the-shelf method uses the same assay for all patients. Without the need of patient specific reagents, it shares the low TAT of a tumor-naïve approach and offers a much simpler logistics than the bespoke method. The challenge is generating an off-the-shelf assay that covers enough of the genome at a low enough error rate (*20–23*). Pre-designed MRD panels targeting cancer-related genes typically use UMI with deep sequencing to achieve high accuracy in variant calling, but the number of markers these panels track for each patient are very limited (*20, 21, 23*). For example, a 130 kb panel covering 139 critical lung cancer-related genes only captures a median of 2 mutations per patient (range: 1–8 mutations) (*21*).

WGS assays have recently emerged as an innovative approach for cancer screening (*11, 24, 25*) and MRD detection (*26–28*). Tumor-informed WGS MRD assays use genome breadth to supplement sequencing depth for sensitivity, overcoming the limitation of input sample amount. However, the standard UMI error correction, which relies on having multiple reads per input molecule, would be cost prohibitive on a WGS scale (*29–31*). Zviran et al. used a read-centric Support Vector Machine (SVM) model to reduce the error rate for WGS somatic single-nucleotide variants (SNV) to 4.96 x 10^−5^ (*32*). By capitalizing on the cumulative signal of thousands of somatic mutations observed in the tumor genome, they reported longitudinal sensitivity of 91% at 92% specificity in bladder cancer using accumulated ctDNA status (*27*). Other WGS technologies using duplex sequencing have demonstrated ultra-low error rate at < 10^-7^ level, however, these methods suffer from low conversion rates, making a low LOD difficult to achieve (*33–35*). There is a need for an efficient and cost-effective genome-wide error correction method to enable WGS for MRD detection with high sensitivity and specificity.

DNA concatemers generated via rolling circle amplification (RCA) of cfDNA physically link replicated copies, allowing error correction at single read level (*36*). The combination of RCA with repeat confirmation eliminates both PCR and sequencing errors. When adapted for targeted sequencing through hybrid-capture or amplicon workflows, concatemer sequencing has demonstrated high conversion rate and sensitivity for liquid biopsy applications including therapy selection and cancer screening (*37, 38*). In this study, we present AccuScan, a WGS solution for ctDNA detection that utilizes concatemer sequencing for genome wide single-read error suppression, enabling fast and sensitive MRD detection and monitoring in cancer patient plasma samples.

## RESULTS

### AccuScan’s genome wide error suppression enables detection of ultra-low ctDNA levels

The AccuScan assay workflow (Fig. 1A) is optimized for low input cfDNA, efficiently capturing double stranded, single stranded and nicked DNA(*38*). cfDNA is denatured and circularized through ligation, followed by whole-genome amplification using RCA, which generates concatemer molecules containing tandem copies of the original template. These concatemer products are sequenced using paired-end 150 bp (PE150) read length and aligned to the human reference genome. Sequences of each copy within a read pair are compared. A change from the reference that is consistent in all copies is a presumed variant; and a change that is inconsistent is likely to be a PCR or sequencing error and is removed. To assess the efficiency of error correction by AccuScan, we sequenced the cfDNA samples from healthy donors (N=3) using both regular WGS and AccuScan WGS (Fig. 1B). The observed average error rate was 9.4×10^-4^ for regular WGS without filtering; The error rate was reduced to 2.8×10^-5^ when requiring overlap and concordance between read pairs. AccuScan with concatemer error correction had an average error rate of 4.2x 10^-7^, which is ∼2,000-fold lower than the unfiltered WGS data, and ∼67-fold reduction when compared to the read pair corrected WGS (Fig. 1B).

**Fig. 1:**
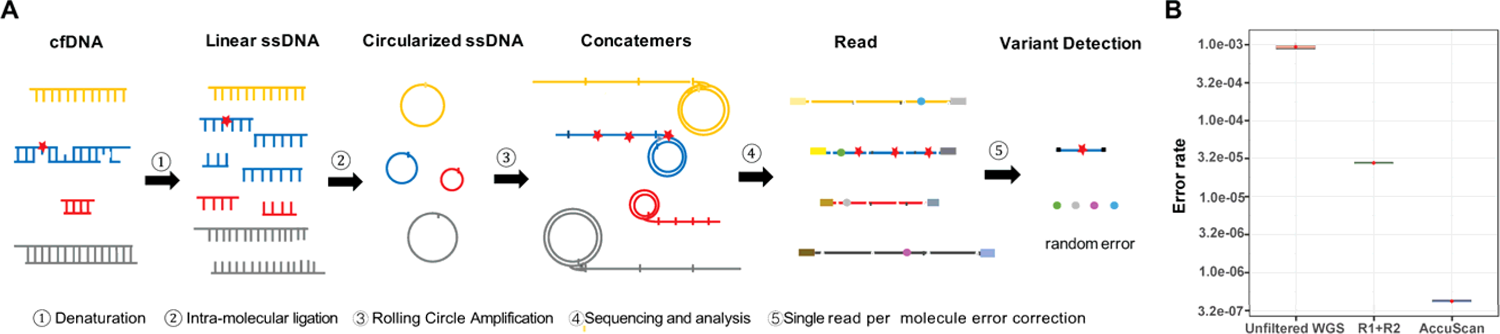
AccuScan: WGS through concatemer error correction. **(A)** AccuScan workflow. Cell-free DNA (cfDNA) is denatured and circularized, followed by rolling circle amplification using random primers to create concatemers. These concatemer products are sequenced and aligned to the human reference genome. A non-reference base that is consistent across all repeats will be called as a mutation. Random errors that are inconsistent between repeats are removed (**B)** Error rate of WGS on healthy human cfDNA samples (N=3) measured by unfiltered reads, read1 and read2 corrected reads and AccuScan. ssDNA, single-stranded DNA; WGS, whole genome sequencing; R1+R2, variants called by requiring read1 and read2 overlap and concordance.

We performed simulations to understand the impact of error rate on ctDNA detection under different cTAF, sequencing depth and number of markers (Fig. 2, fig. S1). A statistical model that calculates the probability of observing expected variants at specific marker loci is used to predict the presence of ctDNA. Sensitivity was calculated as the number of positives predicted over the total number of simulations for each condition, under the nominal specificity setting of 99%.

**Fig. 2.**
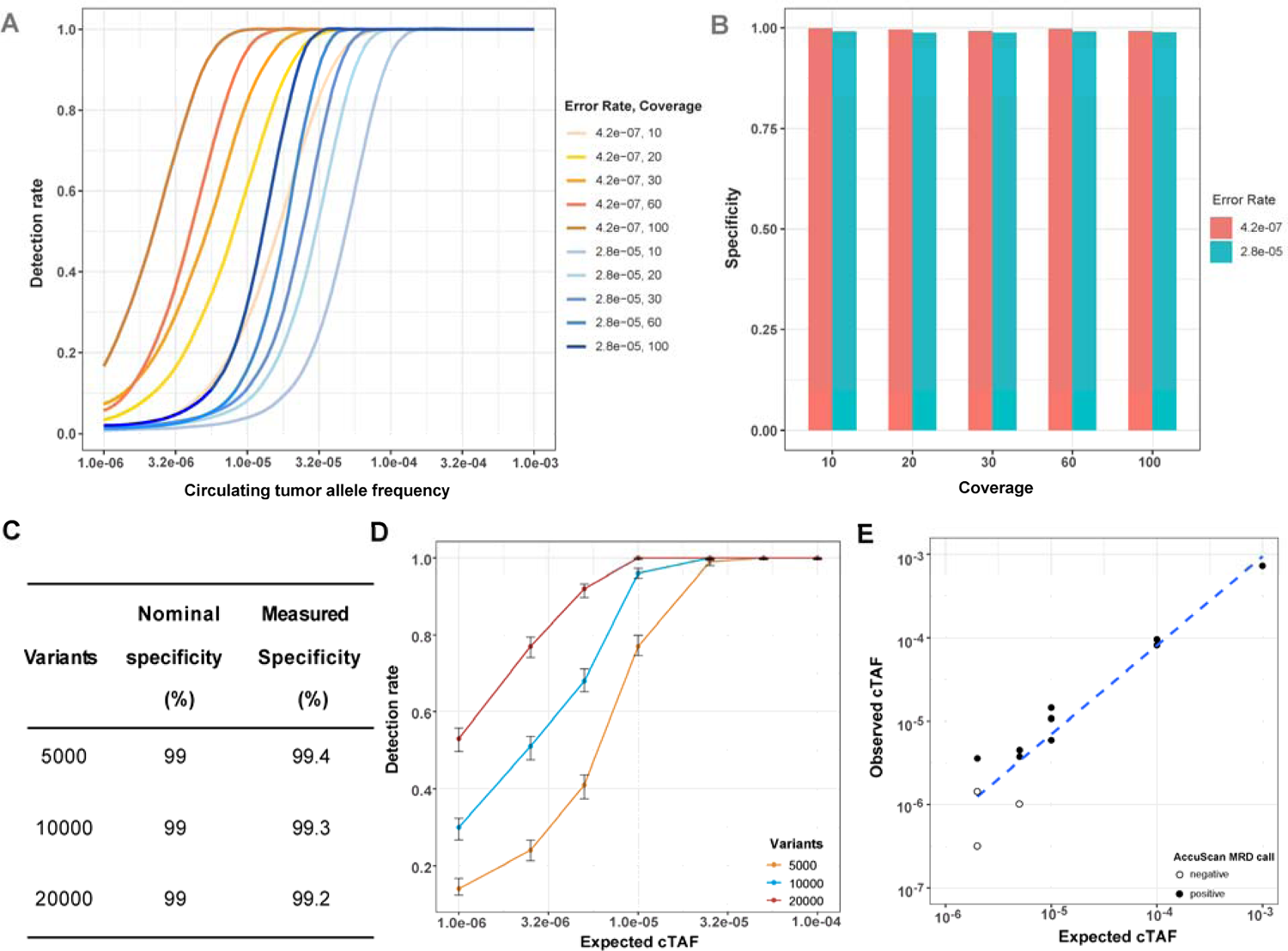
Analytical sensitivity and specificity of AccuScan. **(A)** Simulation using 10,000 markers and two error rates to predict the theoretical detection rates under different sequencing coverages as a function of circulating tumor allele frequency (cTAF). Detection rate is calculated as the fraction of test that are called molecular residual disease (MRD) positive with the nominal specificity set at 99%. **(B)** Simulation using 10,000 markers and two error rates to predict the theoretical specificity with the nominal specificity setting at 99%. Specificity is calculated as the fraction of tests that are called MRD negative when cTAF is 0. **(C-D)** Observed analytical specificity and sensitivity with serial dilutions of healthy cfDNA mixtures simulating cTAF from 1×10^-3^ to 1×10^-6^. Cell-free DNA (cfDNA) from three healthy donors was titrated independently into cfDNA from a different healthy “background” donor. Samples were processed by AccuScan with 10ng input, and tested for MRD by sampling 5000, 10000 or 20000 Single Nucleotide Polymorphisms, 1000 times each, as tumor specific markers. **(E)** Observed cTAF in serial dilutions of a melanoma cfDNA sample with expected cTAF from 1×10^-3^ to 2×10^-6^. Experiments were performed with 1 test at 1×10^-3^, two replicates at 1×10^-4^, 3 replicates at 1×10^-5^, 5×10^-6^, 2×10^-6^ cTAFs.

Figure 2A shows the sensitivity from simulations using 10,000 markers with either 2.8×10^-5^ or 4.2x 10^-7^ error rate under sequencing depth ranging from 10× to 100×. Decreasing error rate or increasing sequencing depth both improved the detection rate. For example, with an error rate of 4.2x 10^-7^ and a 20× sequencing depth, there is a 96% detection rate at a cTAF of 2.5×10^-5^, but when the error rate is 2.8×10^-5^, 100× sequencing depth is required to achieve a similar detection rate at the same cTAF. The specificity with cTAF set to 0 remains above 98.8% across all conditions, which is consistent with the nominal specificity setting (Fig. 2B).

We then measured the analytical performance of AccuScan using healthy sample mixtures. cfDNA from three different healthy “test” donors was titrated independently into cfDNA from a different healthy “background” donor at 7 different concentrations ranging from 1×10^-4^ to 1×10^-6^. These 21 cfDNA mix samples were then sequenced to 60× using AccuScan with 10ng input DNA per reaction and we assessed our ability to detect the “test” donor SNPs from background. Out of the over 100,000 SNVs at which each test and background sample pair differed (table S1), we tested for randomly selected subsets of 5,000, 10,000, or 20,000 SNVs (SNVs were selected to have a variant-type profile similar to CRC tumors). Marker selection was repeated 1,000 times per condition and MRD testing was run with 99% nominal specificity. The observed specificities were ≥99% for 5,000, 10,000 or 20,000 markers conditions (Fig. 2C). The observed sensitivity at 2.5×10^-5^ cTAF and above level was greater than 99% for all conditions tested. At 10 PPM corresponding to 1×10^-5^ cTAF, the average detection rate was 77% for 5,000 markers, 96% for 10,000 markers, and 100% for 20,000 markers (Fig. 2D).

The analytical sensitivity of AccuScan was further confirmed by mixing cfDNA from a melanoma patient with cfDNA from a healthy donor (fig. S2A). The original cancer cfDNA sample had a cTAF of 1.1% as measured by ddPCR of a BRAF V600E mutation present in the primary tumor (fig. S2B). Dilutions were made of 5 different expected frequencies from 1×10^-3^ to 2×10^-6^ and ddPCR of BRAF V600E was performed to confirm the 1×10^-3^ dilution (fig. S2C). The diluted cancer samples were sequenced by AccuScan with 10ng input per reaction. The observed detection rate is 100% for samples with cTAF of 1×10^-3^, 1×10^-4^ and 1×10^-5^, 67% (2/3) at cTAF of 5×10^-6^ and 33% (1/3) at cTAF of 2×10^-6^ (Fig. 2E). AccuScan sequencing of a negative control (cfDNA from a healthy donor) was negative in both replicates.

To further confirm the sample level specificity of AccuScan, we collected more than 1.3M tumor-specific variants from 57 different cancer patients, including colorectal cancer (CRC), esophageal squamous cell carcinoma (ESCC) and melanoma, randomly sampled a subset of 5K, 10K or 20K equivalent variants for testing the MRD call in mismatched patient plasma samples. We did 2,000 random samplings of mismatched variants for each combination of variant count level and plasma sample. The average sample level specificity is computed as the fraction of MRD tests that are characterized with a negative MRD call. The observed values were similar to the nominal specificity, with 99.3%, 99.1%, and 98.9%, for 5K, 10K and 20K variant count levels, respectively. These results suggest that the AccuScan assay and analysis have the intended performance for patient plasmas using tumor variants (Data not shown).

### Identification of tumor-specific variants using a white blood cell free workflow

A tumor-informed MRD test uses tumor-specific variants as markers for tracking the disease. Sequencing of tumor tissues finds not only cancer mutations, but also germline SNPs and other types of variants such as clonal hematopoiesis of indeterminate potential (CHIP) variants. If not removed properly, these non-cancer variants can lead to false positives in an MRD test. One common strategy for filtering non-cancer variants is to remove variants found in the matching white blood cell (WBC) from the same patient. However, this method requires extra sample processing and sequencing (Fig. 3A). To simplify the MRD workflow, we investigated the effect of skipping WBC sequencing and using SNV information from the post-treatment plasma samples to remove germline and CHIP variants (Fig. 3B).

**Fig. 3.**
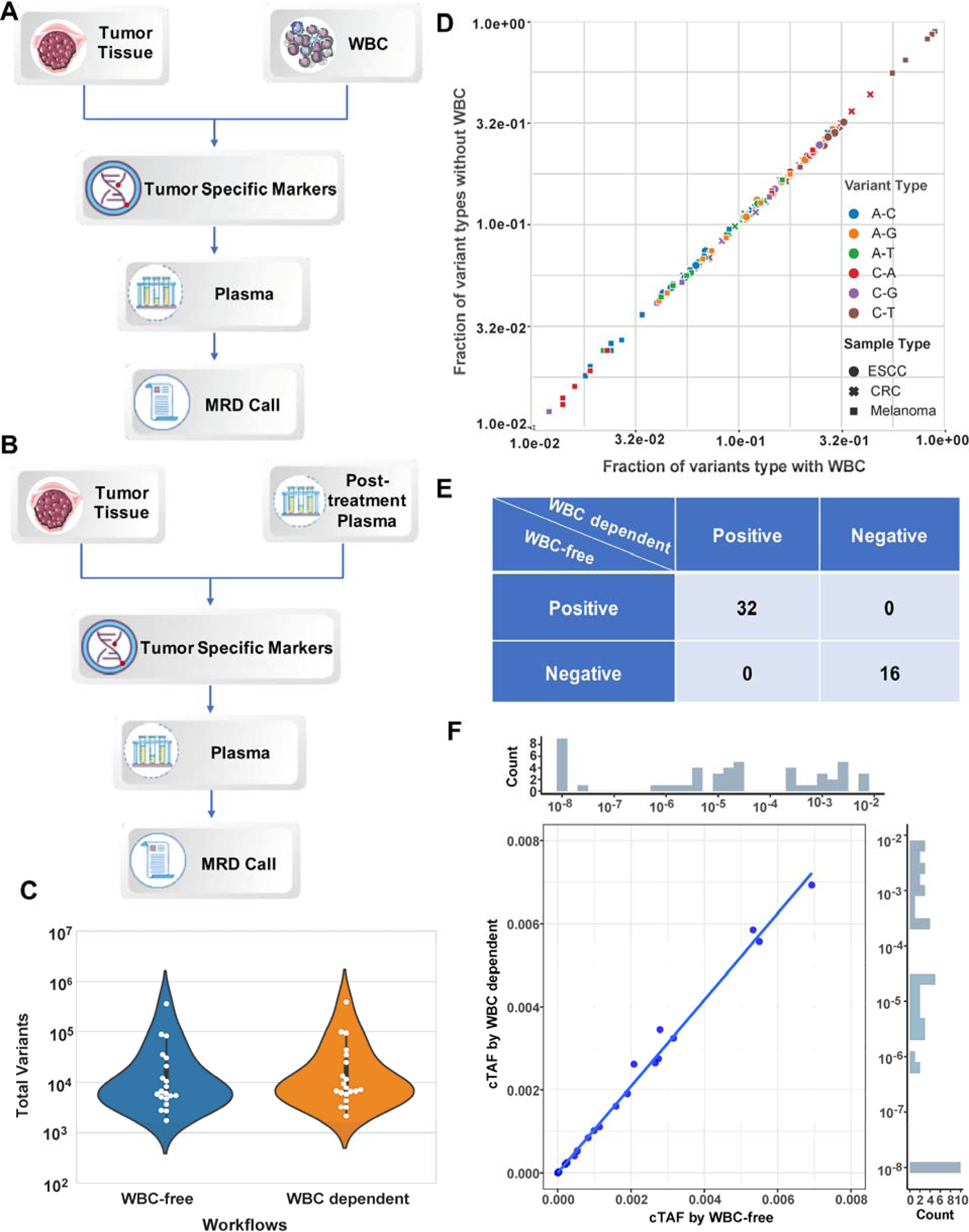
WBC-dependent and WBC-free workflows for AccuScan MRD detection. **(A)** the white blood cell (WBC) dependent workflow uses whole genome sequencing (WGS) data from WBC to remove germline and hematopoiesis of indeterminate potential (CHIP) variants found in the tumor tissue, keeping tumor-specific markers for molecular residual disease (MRD) test of the plasma samples. **(B)** the WBC-free workflow uses WGS data from the low-tumor burden post-treatment plasma samples to remove germline and CHIP variants from the tumor tissue. Any tumor variants that are found in the plasma samples with more than one cell-free DNA molecules are removed from the tumor variant list. The remaining variants from the tumor tissue WGS are used as tumor specific markers for MRD test in plasma samples. **(C)** Total number of tumor specific markers identified with and without WBC sequencing. **(D)** Comparison of the variant profile for tumor specific markers identified using two workflows. **(E)** Comparison of the observed circulating tumor allele frequency (cTAF) in the same plasma samples using tumor specific markers identified by two workflows. **(F)** Comparison of the AccuScan MRD call in the same plasma samples using tumor specific markers identified by two workflows. CRC, colorectal cancer; ESCC esophageal squamous cell carcinoma.

With 40-60× sequencing of a low-tumor-burden (cTAF < 0.1%) plasma sample, germline and CHIP mutations can be found at ≥ 2 molecules level, while tumor-specific variants will generally be found at single molecule level. Hence, we may remove variants found with 2 or more molecules in the post-treatment plasmas from the tumor tissue sequencing result to obtain the list of tumor-specific mutations. To test the feasibility of this approach, we compared the performance of a tumor-WBC workflow with a WBC-free workflow using matched tumor tissue, WBC and plasma samples collected from 20 cancer patient samples (table S2). The number of tumor specific variants and variant type profiles found by the two different workflows are shown in Figure 3C-D. Overall, the number of mutations identified by both methods were very similar, as were the variant-type profile of the mutations identified. AccuScan MRD analysis of the plasma samples (n=48) from these 20 patients returned identical MRD calls under either workflow (Fig. 3E), and the cTAF values were strongly correlated (R^2^=0.99, Fig. 3F). These results suggest that post-treatment plasma can be used in the place of WBC for tumor-specific variant identification.

### MRD detection and prognostic value in surgical patients

We next evaluated the performance of AccuScan for MRD detection in post-surgical gastrointestinal cancers, including 32 CRC patients and 17 ESCC patients.

The CRC patients were at diverse clinical stages (22% stage I, 38% stage II, 34% stage III, 6% stage IV) and received radical surgery (table S3). Formalin Fixed Paraffin Embedded (FFPE) samples were available from all patients. For the 15 patients with available WBC, we used the tumor-WBC workflow to identify tumor-specific variants; for all other patients, we used the first post-operative (post-OP) plasma samples for the WBC-free workflow (Fig. 3B; table S2). We identified a median of 5,820 tumor-specific variants per patient (2148-265800, Fig. 4A), which correspond to ∼2 mutations/Mb.

**Fig. 4.**
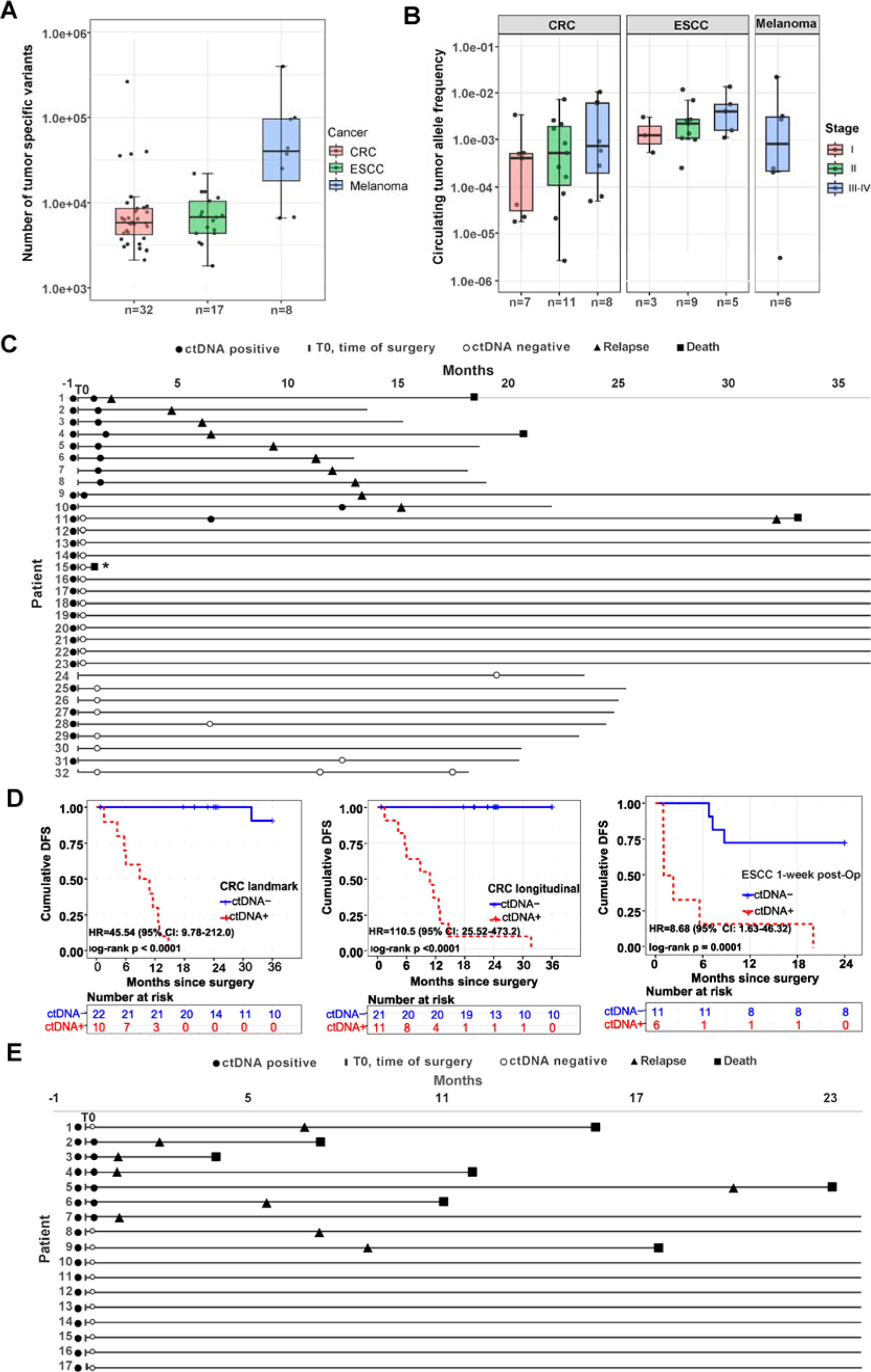
AccuScan molecular residual disease detection in clinical samples. **(A)** Number of tumor specific variants identified in CRC, ESCC and melanoma patients. **(B)** Observed circulating tumor allele frequency (cTAF) in pre-treatment plasma samples. **(C)** Swimmer plot of CRC patients who undergo radical surgery. *The patient died due to non-cancer related reasons. **(D)** Kaplan-Meier disease-free survival analysis (DFS) for CRC surgical patients using blood samples collected at landmark time point only or longitudinal samples collected overtime, and for ESCC surgical patients using blood samples collected within 1 week after surgery. Patients who are circulating tumor DNA (ctDNA) positive in the post-operative (post-OP) plasma samples showed significantly shorter DFS. **(E)** Swimmer plot of ESCC patients. All patients received curative surgery. Patient blood samples were collected before surgery and 1 week after surgery. CRC, colorectal cancer; ESCC esophageal squamous cell carcinoma; ctDNA, circulating tumor DNA; ctDNA+, ctDNA positive; ctDNA-, ctDNA negative.

Of the 32 patients, 26 had plasma samples collected before surgery and 28 had plasma samples collected at landmark (within one month of surgery). There were 7 additional plasma samples collected at time points later than one month after surgery. The median follow-up time in this CRC cohort was 24.13 months (IQR: 18.5-36). ctDNA was detected in all the 26 pre-operative (pre-Op) cfDNA samples with a median cTAF of 5.2×10^-4^ [interquartile range (IQR): 6.6×10^-5^-2.4×10^-3^] (Fig. 4B). 34.4% (11/32) of patients had ctDNA detected in the post-Op samples. All patients that are ctDNA positive in the post-Op samples relapsed within 3 years after surgery (Fig. 4C, 4D). The post-Op ctDNA positive patients had shorter DFS times than ctDNA negative patients (HR, 45.54, 95%CI: 9.78-212, log-rank p< 0.0001) (Fig. 4D). The median disease-free survival (DFS) of the ctDNA positive patient group was 10.8 months (IQR: 5.8-12.7), with 63.64% (7/11) of ctDNA positive patients had a recurrence within one year, and 90.91% (10/11) of ctDNA positive patients relapsed within two years. One of the ctDNA positive patients, patient #11, was ctDNA negative at the first landmark timepoint, converted to ctDNA positive at 6 months post-Op and then relapsed at 32 months. Patients that were ctDNA negative at all post-Op time points were progression free during the follow-up period (up to 36 months) (Fig. 4C). Taken together, these results suggest 90% (95% CI: 55.5%-99.8%) sensitivity at landmark, 100% sensitivity with longitudinal monitoring, 100% (95% CI: 80.5%-100%) specificity, and 96.3% (95% CI: 81%-99.9%) accuracy for predicting CRC recurrence.

The ESCC cohort included patients from stage I-III (18% stage I, 53% stage II, 29% stage III) and received curative-intent surgery (table S4). FFPE, WBC, pre-Op plasma, and very early post-Op plasma samples were collected from all patients. The post-Op plasma samples were collected right before the patients were discharged from the hospital, which is 1-week after surgery. Using the paired tumor and WBC samples, we identified a median of 6,768 tumor-specific variants per patient (Fig. 4A).

The follow-up time of this ESCC cohort ranged from 4.03 months to 24 months. ctDNA was detected in all 17 of pre-Op samples with a median cTAF of 0.27% (IQR: 0.13%-0.55%) (Fig. 4B). In the post-OP plasma samples, ctDNA was detected in 35.29% (6/17) of the patients, with a median cTAF of 1.3×10^-4^ (IQR: 1.9 x 10^-5^-1.1 x 10^-2^). ctDNA positive patients had shorter DFS times than ctDNA negative patients (hazard ratio, HR, 8.68, 95%CI: 1.63-46.32, log-rank p=0.0001) (Fig. 4D). All the patients with ctDNA positive post-op samples (6/6, 100%) had a disease recurrence within 2 years of surgery; 5 of 6 (83%) patients had a recurrence within one year (Fig. 4E). In contrast, of the 11 patients with ctDNA negative post-OP sample, 8 patients remained disease-free patients were followed for 24 months, and 3 patients relapsed. ctDNA detection at 1-week post-Op had 66.67% (95% CI: 29.93%-92.51%) sensitivity, 100% (95% CI: 63.06%-100%) specificity and 82.35% (95% CI: 56.57%-96.27%) accuracy in predicting ESCC recurrence.

### ctDNA monitoring during immunotherapy

Advances in immune checkpoint blockade (ICB) have significantly improved survival of patients with advanced melanoma (*39*). However, only a fraction of patients (<20%) respond to ICB. There is an urgent need for means of prognosis and monitoring of patients undergoing immunotherapy. We explored the potential use of AccuScan for immunotherapy monitoring in a pilot study with advanced melanoma. A total of 22 plasma samples were collected from 8 melanoma patients, including 6 samples collected before any treatment (table S5). WGS of the paired tumor and WBC DNA samples identified a median of 34,323 SNVs, with an average of 90,006 tumor specific SNV per patient (Fig. 4A). All 6 pre-treatment samples were ctDNA positive with cTAF measured as low as 2.82×10^-6^ (Fig. 4B, table S6).

Of the 8 melanoma patients, 6 had at least two plasma samples collected during ICB treatment. We therefore measured ctDNA kinetics and compared the results with radiological data in these 6 patients. For patients 1 through 5, radiographic changes matched the AccuScan measured cTAF changes (Fig. 5, table S6). Patient 1 showed rapid decline of cTAF from baseline to C2, and clearance of ctDNA at C4 time point. Patients 2 was ctDNA positive before surgery, converted to ctDNA negative after and maintained ctDNA negative by C3 of adjuvant ICB therapy. Both patients had sustained complete response with no disease recurrence through the monitoring period. Patients 3 and 4 had very high cTAF levels in all plasma samples. For Patient 3, cTAF increased from 12.5% at baseline to 16% before C2 and computed tomography (CT) scan detected disease progression two months later. Patient 4 observed tumor regression by imaging at 2.6 month after C1, while AccuScan test suggested cTAF increased to ∼0.7% at 4-month timepoint, and 2.5 month later, CT scan confirmed tumor progression. Patient 5 was ctDNA positive with persistently low cTAF (∼1×10^-5^) in plasma samples taken before and during adjuvant ICB treatment, which is consistent with CT scans showing stable lung nodules.

**Fig. 5.**
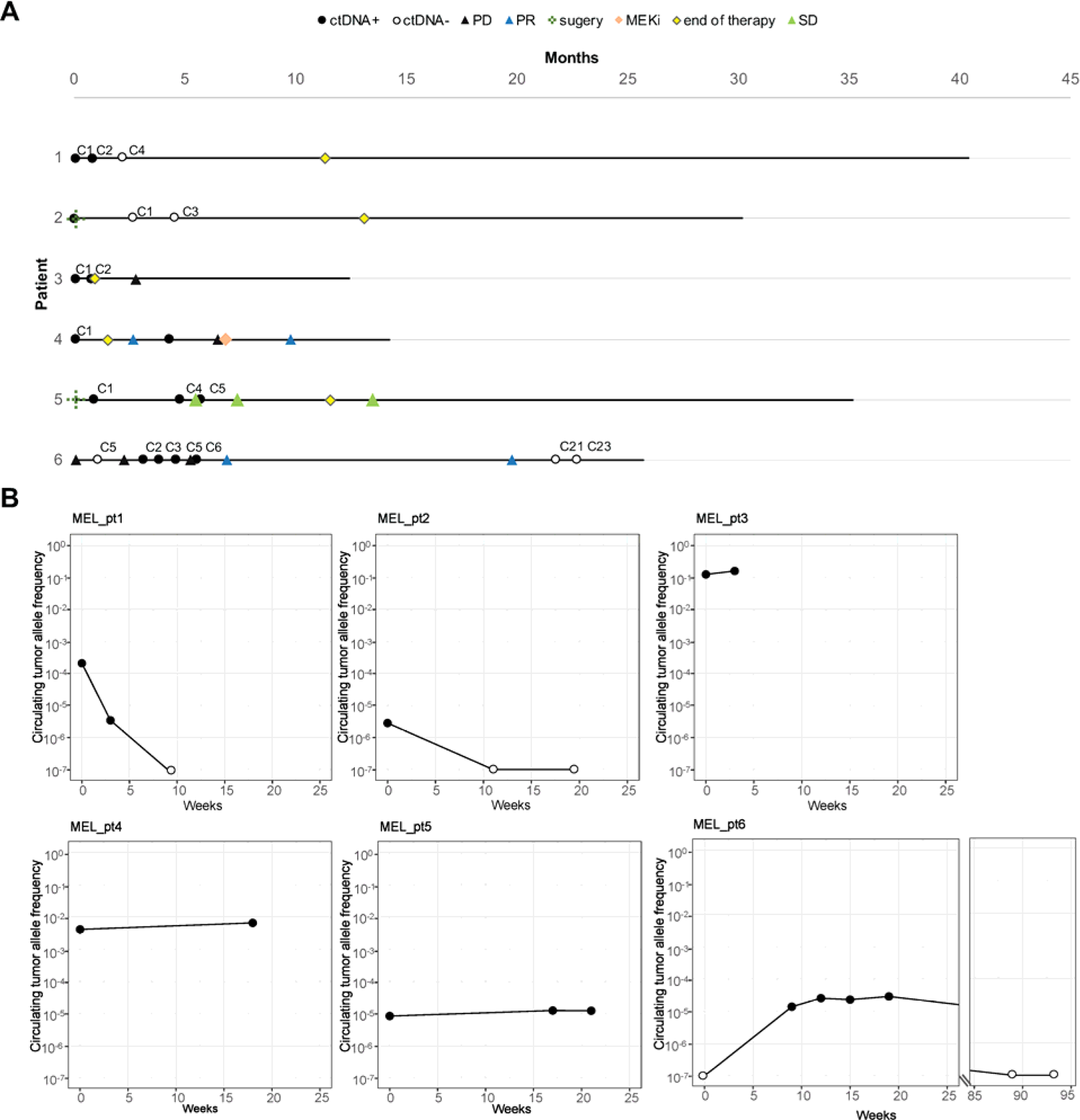
AccuScan for treatment monitoring of melanoma patients. **(A)** Swimmer plot of melanoma patients who undergo surgery and immunotherapy. **(B)** Dynamic change of circulating tumor DNA level in melanoma patients over time. ctDNA, circulating tumor DNA; C, cycle, eg. C2 for cycle 2.

Patient 6 did not have a pre-treatment sample. The first plasma sample was collected after 4 cycles of adjuvant therapy with nivolumab, followed by 6 samples collected during ipilimumab/nivolumab therapy (Fig. 5A). Based on the MRD model, the first sampling timepoint was ctDNA negative with cTAF below 3.8×10^-6^ (95% confidence interval, table S6), while CT scan detected a 4mm lung nodule. The cTAF elevated to 1.4×10^-5^ at the second blood-draw (table S6), which was consistent with the increased tumor burden measured by the second CT scan. ctDNA level stabilized after the third blood test, while the imaging data showed continuous tumor progression. The fourth scan, which was taken 1.5 months after the fifth ctDNA test, showed excellent partial response, and the patient reached near complete response (CR) and ctDNA clearance 13 months later (Fig. 5B). These results indicate that the observed progression by third imaging was likely to be pseudo-progression, and the early stabilization of ctDNA level may signal that patient was responding to immunotherapy.

In summary, a total of 117 plasma samples from 57 cancer patients were processed in this study. Of all the cancer plasma samples analyzed in this study, 96.6% (113/117) samples had total cfDNA amount <60ng, and 17.1% (20/117) of samples had total cfDNA < 10ng (table S7, fig. S3). Of the cancer plasma samples that are tested ctDNA positive, about 30% are with cTAF <1×10-4, and ∼9% are below 10 PPM (fig. S4).

## DISCUSSION

This is the first report of applying genome-wide concatemer error correction to tumor-informed MRD detection and monitoring in cancer patients. AccuScan combines RCA and linked reads to remove artifacts introduced during library preparation and sequencing, demonstrating highly efficient and accurate measurement of ctDNA in both contrived samples and clinical samples.

Scarcity of ctDNA remains as the major challenge for MRD detection. As most of the cfDNA molecules are from WBCs, the fraction of tumor-derived DNA fragments can be significantly lower than 0.01% in the post-treatment patient plasma samples (*40*). Detecting tumor specific DNA at such low frequency requires techniques with high sensitivity and specificity. Tumor-naïve MRD tests using epigenetic signals showed moderate sensitivity at cTAF >10^-4^ level. The sensitivity of ctDNA detection can be significantly improved with tumor-informed MRD assays, by either sequencing all the molecules exhaustively at selected genomic loci or tracking large number of tumor-specific mutation markers with relatively shallow depth. The first approach often requires personalized reagents, which increased logistical challenges and TAT. In addition, it requires high input DNA amount, such as >60ng, which may not be always available in clinical settings (*15*). With the drop of sequencing price, WGS has emerged as an attractive solution for MRD given its relatively simple workflow and low sample input requirement. A WGS approach allows tumor and plasma samples to be sequenced in parallel, enabling fast turnaround. The amount of DNA a WGS test requires can be 10ng or lower. When combined with a home-blood collection device, it may greatly simplify logistics and improve patient experience.

The sensitivity of a WGS-based MRD test is constrained by the WGS error rate. Simulation data showed that a WGS test with a reduced error rate can achieve a lower LOD; and for the same LOD, a test with fewer errors requires significantly fewer reads compared to a WGS test with a higher error rate (fig. S1). AccuScan reduces the SNV error rate to less than 1 in 2 million with PE150 sequencing, enabling LOD95 of 5×10^-5^ at 10× coverage with 10,000 variants, and detection of tumor specific mutations down to ≤10 PPM at 60×, while maintaining a high sample-level specificity of 99%. Comparable analytical sensitivity was achieved with contrived samples, including mixtures of normal cfDNA and dilutions of tumor plasma into normal cfDNA. The AccuScan error rate can be further improved with longer sequencing reads. Under PE150 sequencing, the R1 and R2 sequences of a short library molecule with only one copy of the cfDNA can be mistakenly treated as two independent repeats for variant confirmation. This may lead to some of the AccuScan residual errors, and such errors can be readily identified and removed by long reads. Preliminary data showed that single end 300 base sequencing further lowered the error rate to 3.1×10^-7^ (fig. S5), suggesting that longer reads can effectively clean up residual errors by ensuring correction with 2 or more copies of independent repeats.

The sequencing depth needed for a given sensitivity requirement is inversely correlated with the number of mutations for a WGS-based MRD assay. The average mutation rate, which meters the average WGS depth needed for MRD, is ∼4.0 mutations/Mb measured across cancer types (*41–43*). Relatively low mutation rate was observed in pediatric malignancies (median 1.7 mutations/Mb), while disease associated with mutagen exposure such as lung cancer or melanoma showed high mutation rate (median mutation rate 7.2 mutations/Mb and 13.5 mutations/Mb, respectively) (*42*). In our study, we observed the average mutation rates of ∼2.6 mutations/Mb, 5.6 mutations/Mb and 26.7 mutations/Mb for ESCC, CRC and melanoma respectively. Given such mutation rates, AccuScan can reach 10 PPM or lower with an average of 60× sequencing coverage.

In clinical practice, the sensitivity and specificity requirements for a MRD test depend on the specific applications. AccuScan sequencing depth can be adjusted based on cancer types and mutation rate of the patient to achieve an optimal cost-benefit balance while accommodating different application needs. For example, National Comprehensive Cancer Network guideline recommends starting ACT for high-risk CRC patients no later than 6-8 weeks after surgery (*44*). The landmark timepoint, which is within a month of surgery, is key for clinicians to decide between escalation or de-escalation of therapy. Fast TAT and high sensitivity at landmark are critical. They give oncologists the time and confidence to de-escalate treatment while ensuring most, if not all, high risk patients receive needed adjuvant therapy following their surgery. In this study, AccuScan had a landmark sensitivity of 90% to predict relapse after surgery with 100% specificity in CRC patients. Considering an average sensitivity of 45%-56% of landmark sensitivity for CRC with current commercial offerings, AccuScan picked up as much as twice the number of patients who may benefit from adjuvant therapy without increasing over treatment. During longitudinal monitoring for patients who have shown partial response or CR, high specificity is crucial for avoiding anxiety, unnecessary exposure to toxic side effects and financial burden associated with the treatments. Our pilot study of immunotherapy monitoring with melanoma patients suggests that AccuScan measured ctDNA dynamic changes can be combined with imaging for better predicting patient response. Large clinical studies with controlled patient cohorts will be needed to understand how this information can be integrated in the clinical setting for therapy management.

In addition, WGS data offers not only SNV results, but also other rich molecular information including copy number variants, structural variants, tumor mutation burden (TMB), mutation signatures as well as epigenetic information such as fragmentomics. A WGS technology with low error rate like AccuScan would significantly improve the efficiency and accuracy for TMB and tumor mutation signature detection in cfDNA. In addition, studies have shown that using machine learning to integrate genetic information from multiple somatic variant types significantly improved the performance of tumor informed MRD detection (*32*); and the combination of genetic and epigenetic information, such as the SNV and fragmentomics signal, may improve the sensitivity of cancer early detection (*24, 38*). AccuScan as a single strand sequencing technology captures small cfDNA fragments representing regulatory protein binding footprints (*38*). Combining the mutation and fragmentomics signal from AccuScan data renders great potential to build a tumor naïve test for MRD detection and cancer screening.

From the above data, we can see that AccuScan has great potential in a wide range of research and clinical applications, but we must acknowledge that the sample size of this study was insufficient, we need to confirm it in our ongoing larger investigations on CRC and ESCC.

In summary, AccuScan MRD test demonstrated a high success rate for processing cfDNA samples of a wide range of DNA input amount, with the lowest input of less than 5ng (table S7, fig. S3). We observed high sample-level specificity of 99%, with 100% sensitivity in all pre-treatment samples from CRC, ESCC and melanoma, 90% landmark sensitivity for CRC (<4 week after surgery), and 67% sensitivity for ESCC using plasma samples collected within one week after surgery. The current performance of AccuScan is based on a simple statistical model using SNV information only. Its performance can be potentially enhanced through baseline modeling with accumulation of cfDNA WGS data and incorporation of machine learning strategies.

## MATERIALS AND METHODS

### Experimental Design

A total of 58 subjects including 32 CRC patients, 17 ESCC patients, 9 melanoma patients and 4 healthy individuals were included in this study. This study was conducted according to the Declaration of Helsinki and was approved by the Institutional Review Boards (K-KYSB-2021-005, CHEC2020-021, 0609001869). Written informed consent was obtained prior to the initiation of the study. Blood samples were collected in blood collection tubes (BD Vacutainer K2EDTA tubes for CRC and ESCC, catalog no. 367525; BD sodium heparin tubes for melanoma patients, catalog no. 367874). FFPE tumor tissues were collected from resected CRC and ESCC biopsies. Fresh frozen tumor tissues were collected from melanoma tumor biopsies.

### Plasma DNA processing

Blood samples were centrifuged within 4 hours after blood drawing. For ESCC and CRC, plasma sample was separated by two steps: 1) 10 min at 1,900 g at 4 °C, WBCs were collected from the middle phase and stored at –80 °C;2) the upper phase of the first centrifugation was collected and centrifuged for another 10 min at 16,000 g at 4°C. cfDNA extraction procedures were in accordance with the manufacturer’s instructions. Isolation of cfDNA from 1 ∼ 2 mL of plasma was performed using MiniMax High Efficiency Cell-Free Isolation Kit (catalog no. A17622CN-384, Apostle) and eluted in 80 µL Tris-EDTA buffer. The extracted cfDNA was qualified by Qubit fluorometer 3.0 (catalog no. Q33216, Thermo Fisher Scientific) and 2100 bioanalyzer (catalog no. G2939BA, Agilent).

For melanoma patient samples, plasma was collected after spinning the tubes at 800xg for 10 minutes. The collected plasma is then spun again at 450xg for 10 minutes before being aliquoted and stored at –80 °C. PBMC are isolated using lymphoprep (catalog no. 07801_c, STEMCELL Technology).

### Tissue Genomic DNA Processing

For CRC and ESCC patients, The TIANamp Genome DNA Kit (catalog no. DP304-03, TIANGEN) was used to extract gDNA of WBCs and frozen tissue. gDNA was extracted from paraffin sections of tumor samples by High Pure FFPET DNA Isolation Kit (catalog no. 6650767001, Roche). The melanoma tumor and matching WBC DNA were extracted from fresh frozen samples with Qiagen DNeasy Blood and Tissue Kit (catalog no. 69506, QIAGEN).

### Library preparation and sequencing

gDNA of WBCs and tissue samples were sonicated into short fragments with a peak around 300 bp by M220 ultrasonicator (catalog no. 500295, Covaris), 100 ng fragmented gDNA was used for library construction using KAPA Hyper Prep Kit (catalog no. KK8504, KAPA) following the manufacturer’s protocol. The gDNA libraries were sequenced on DNBSEQ-T7 (MGI) with paired-end 150 bp mode. cfDNA library preparation was performed as described previously (*38, 45*). Briefly, cfDNA molecules were denatured and circularized by intra-molecular ligation, followed by rolling circle amplification (RCA). The whole genome RCA products were sonicated into fragments with a peak at around 500 bp by ultrasonicator, followed by library construction using NEBNext® Ultra™ II Ligation Module (catalog no. E7595L, NEB) and NEBNext® Ultra™ II End Repair/dA-Tailing Module (catalog no. E7546L, NEB). The cfDNA libraries were sequenced on DNBSEQ-T7 (MGI) with DNBSEQ-T7RS High-throughput Sequencing Set (FCL PE150).

### Tissue variant pipeline and tissue-informed marker selection algorithm

WGS reads were demultiplex using Complete Genomics sequencer software and processed by Sentieon® TNseq® for variants calling. The tumor-normal mode was used when WBC data was available; otherwise, the tumor-only mode was used. When WBC data was not available, we removed potential germline and CHIP variants by filtering out any variant found with more than one unique molecule in a post-treatment plasma sample (which is expected to have a very low tumor burden). C to T substitutions at CpG sites were removed from the variant list for MRD analysis.

### AccuScan error correction algorithm and error rate measurement

Variant calls using concatemer has been previously described in (*45*). Summarizing, reads were aligned to the reference genome (hg38) and high read quality single nucleotide substitutions different to the reference are further investigated for repeat confirmation. Differences from the reference that were consistently supported within a read-pair by at least two tandem copies were considered as true variants; inconsistent differences were considered as random errors and discarded.

For computational reasons, error rates were calculated for a set of randomly selected 200Mb positions of the genome. Error rates were calculated for every possible variant in the selected loci, except for C to T variants at CpG sites, which were excluded for overall error rate calculation. Each possible single nucleotide variant within this region is annotated, filtered according to the same rules as applied on tissue-informed marker selection, and sorted into variant categories. Within a given plasma sample, we removed variant positions where two or more variant molecules were observed. The error rate was calculated as the sum of all observed variant molecules divided by the total sum of molecules interrogated for each variant type.

### AccuScan MRD algorithm

The relative amount of tumor DNA was estimated using a probabilistic model that considered base calls at a set of target locations. The set of locations was specific to each patient and was derived from the list of somatic variants inferred as originating from tumor cells. The number of tumor base calls was modelled as a random variable that depends on the sequencing depth, error rates, and tumor allele frequency.

We estimate the cTAF by maximizing the likelihood of the observations with respect to the variant allele frequency (vAF, vAF 2c 0) and the error rate. For each observation of a variant site, there is a probability P_l_ of observation of the variant, which we can approximate as Pl ∼vAF +e_l_, where vAF is the variant allele frequency and e_l_ is the error rate for that site and variant. We can combine all variant sites of the same variant type with the error rate e_V_ and compute the number of variant observations as a binomial random variable for that variant type with binomial parameter P_V_, P_V_ ∼v_AF_ + e_V_. We assume that the variant types v∈V are determined by the type of mutation as following:

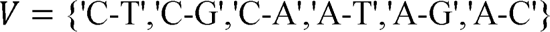

the log-likelihood function for combining independent binomial distributions of all different variant types is

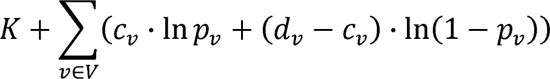

where c_V_ is the number of observations of variant type v, d_V_ is the total depth for that type of variant site, and K does not depend on P_V_ or vAF. We obtain the point estimate for cTAF maximizing the log-likelihood by first finding the derivative (with respect to vAF) and then estimating the value of vAF that makes that derivative 0.

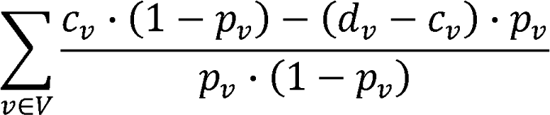

Our null hypothesis is that variant observations are strictly due to errors. To test this hypothesis, we first consider that the null hypothesis corresponds to an extreme of the range of possible values for the parameter (vAF - 0, with vAF 2c 0), then find the difference in log-likelihoods at the point estimate of vAF and at 0, and reject the null hypothesis when the cumulative distribution of a x^2^ distribution with 1 degree of freedom evaluated at the difference in log-likelihoods is greater 0.98. This corresponds to a specificity of 0.99 (99%) for the test (*46*). We obtained 95% confidence bounds for our cTAF estimates: two-tailed confidence intervals for positive samples and one-tailed upper bound for negative samples.

### Simulation of detection rate for different error rates

We simulated theoretical performance given different error assumptions, depths, and tumor fractions. For a given number of 5000, 10000, 20000, 40000 variants and an expected depth in sequencing (10–100), we calculated the chance of seeing a molecule. Each molecule has a probability of being observed as a variant following two binomial distributions, one using the allelic fraction as probability and one using the error rate as probability. We performed 100,000 simulations for each combination of allelic fraction, depth, error rate, and evaluated the outcome using the MRD algorithm described above. Sensitivity was calculated as the number of positive predicted over the total number of simulations for each combination. Specificity was measured at ∼99% using an allelic fraction of 0 as input, as expected from the MRD models parameter settings.

### Analytical Sensitivity and Specificity calculation using contrived samples

For the healthy titration experiment, three “test” cfDNA from three healthy individuals were independently spiked-into one “background” cfDNA samples, to create three sets of serial dilution samples. SNPs for the titration study were selected among autosomal and X-linked SNVs using following criteria: 1) Sites were required to be heterozygous in one individual and absent in the other individuals. 2) Sequencing depth was required to be between 20X and 100X at the variable sites for all individuals. 3) The observed variant allele frequency was between 0.4 and 0.6 for the selected sites. 4) SNPs from problematic regions, as well as C->T CpG variants were excluded. 5) SNPs present in multiple plasma samples were excluded.

This resulted in between 124K and 165K SNPs per sample. These SNPs were used as markers for the presence of the test sample in the dilution. Depth of sequence and presence of the donor allele was assessed at each of the target sites. For each plasma titration sample, we randomly subsampled 5k, 10k, or 20k variants constrained to having the same distribution of mutation types as in a typical CRC tumor. 1000 rounds of subsampling were obtained at each variant level for every plasma titration sample, and the MRD algorithm was run on each of them. Sensitivity was measured as the rate at which subsamples were called as positive. For the specificity analysis, we generated random mutations at locations adjacent to those of the germline variants used in the sensitivity analysis, as long as they did not result in CpG sites. We randomly subsampled 5k, 10k, or 20k variants, also constrained the subsamples to having the same distribution of mutation types as in a typical CRC tumor. The MRD algorithm was run on each of the 1000 subsamples generated for each plasma titration. Specificity was measured as the fraction of subsamples that resulted in a negative MRD call.

For the cancer titration experiment, we diluted a plasma sample from a melanoma cancer patient into a serum sample from a healthy individual to different concentration levels ranging from 1×10^-3^ to 2x 10^-6^. We then sequenced the contrived plasma samples and ran the MRD analysis using the variant list from the melanoma tumor sample.

### AccuScan Clinical Specificity

To evaluate the performance of the assay, we evaluated the specificity of our assay in patient plasmas using tumor variants found in mis-matched patient tumors. Total ∼1.3M tumor (somatic) variants were collected from 57 patients, including the 32 CRC, 17 ESC and 8 Melanoma patients. We imposed all conditions required in tumor-variant-selection. For each plasma sample, we first excluded variants that were observed in the tissue or WBC BAMs of the corresponding patient. In each case we sampled the number of variants depending on the average depth in the plasma sample, so that the expected sum of depth was equivalent to sampling 5k, 10k or 20k variants with expected average depth of 60. For each plasma and equivalent number of variants we generated 2000 random subsamples and ran the MRD test on each one using a specificity of 99%. The fraction of subsamples with a negative value for the MRD test provides the empirical specificity.

### Statistical Analysis

Data visualization and statistical analysis were done using R (4.2.2, The R Foundation for Statistical Computing, Vienna, Austria, https://www.r-project.org/). The two-sided test P < 0.05 shows statistical significance.

## Data Availability

The raw sequencing data generated in this work is not publicly available due to national legislation. Raw Data of this project can be accessed after we getting the authorization from the Ministry of Science and Technology of The People's Republic of China. The datasets used and/or analyzed during the current study are available in the main text or the supplementary materials.

## Acknowledgments

**Funding**: Shanghai Natural Science Foundation Project (21ZR1458200) Key talent introduction project of Tongji Hospital (2021) Clinical Research Incubation Program of Tongji Hospital ITJ(ZD)2104 NIH Yale SPORE in Skin Cancer NCI P50CA121974

## Author contributions

Conceptualization: LW, MF

Methodology: LW, MF, PT, TW, FLM, YYW, AR

Sample and clinical information collection: XXL, TL, MXL, WC, AR, MWB, MS, QY, RH

Investigation: XXL, TL, MXL, WC, AR, MWB, MS, QY, RH

Visualization: QQY, LW

Funding acquisition: ZQH, MWB, MS, QY, RH

Project administration: HJ, ZQH, RH

Writing – original draft: XXL, TL, MXL, LW, MF, PT, TW, FLM, YYW, QQY

Writing – review & editing: XXL, TL, MXL, WC, LW, MF, PT, TW, FLM, YYW, QQY, HJ, ZQH, AR, MWB, MS, QY, RH

## Supplementary Materials

### Supplementary figures

**Fig. S1.**
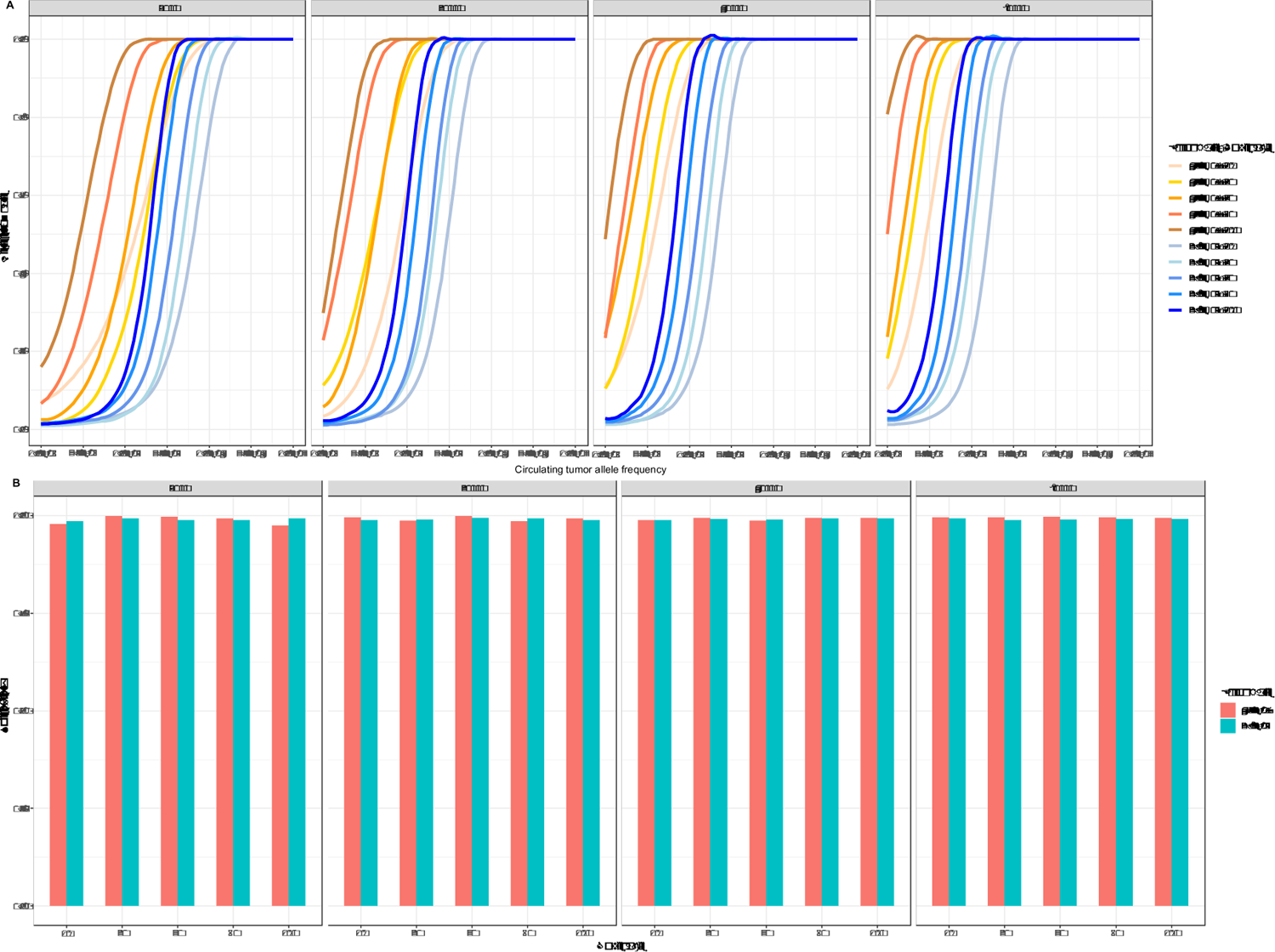
Analytical sensitivity and specificity of AccuScan. **(A)** Simulation using 5000, 20000, 40000, 80000 markers and two different error rates to predict the theoretical detection rate under different sequencing coverages as a function of circulating tumor allele frequency (cTAF). The 4.2x 10^-7^ error rate showed higher sensitivity than the 2.8×10^-5^ error rate under same sequencing depth. Detection rate is calculated as the fraction of tests that are called molecular residual disease (MRD) positive with the nominal specificity set at 99%. **(B)** Simulation using 5000, 20000, 40000, 80000 markers and two different error rates to predict the theoretical specificity with the nominal specificity setting at 99%. Specificity is calculated as the fraction of tests that are called MRD negative when cTAF is 0.

**Fig. S2.**
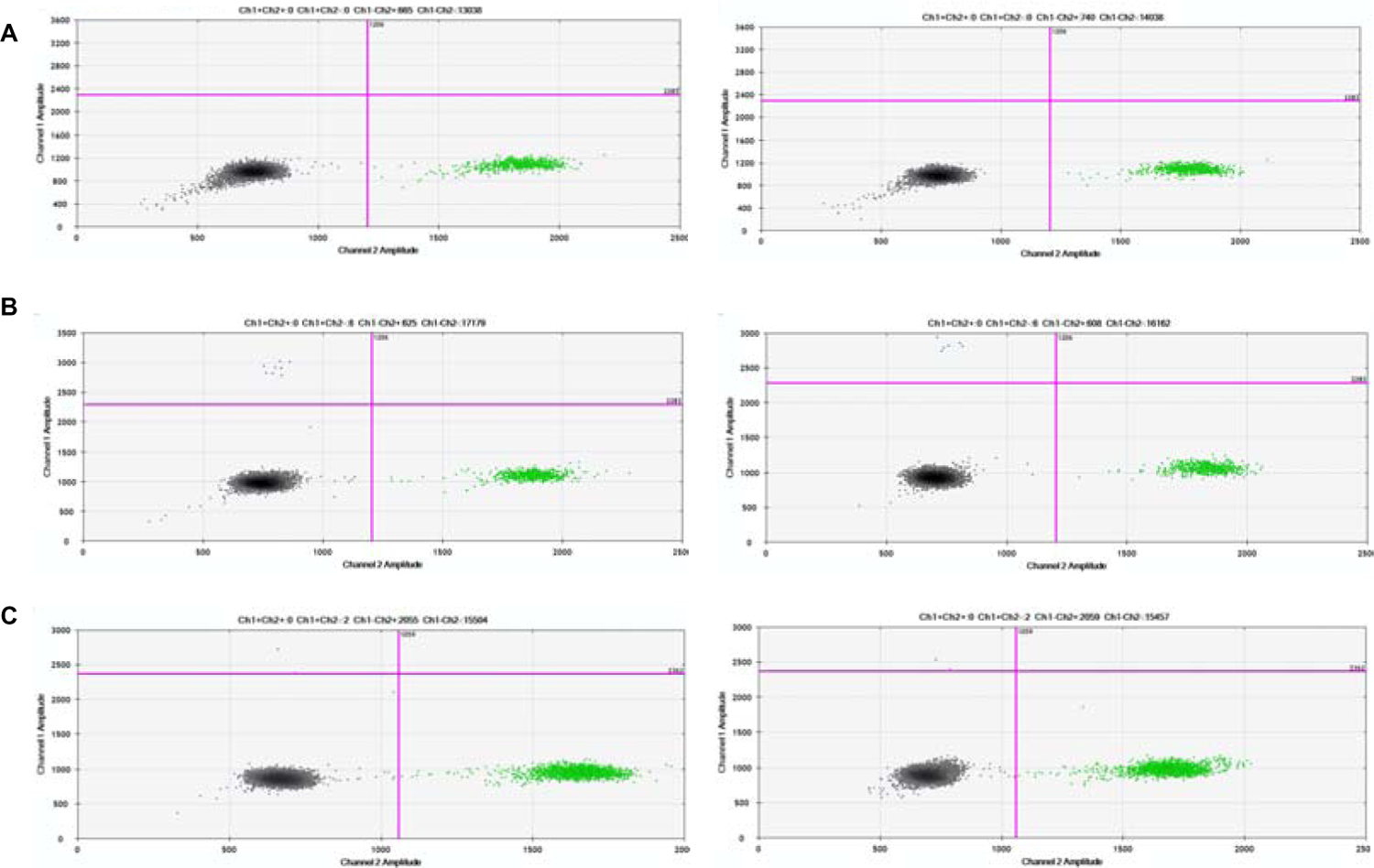
Droplet digital PCR of the melanoma cancer cell-free DNA sample. **(A)** Droplet digital PCR (ddPCR) analysis of a healthy plasma sample; **(B)** ddPCR analysis of the original melanoma cancer cell-free DNA (cfDNA) sample using the BRAF V600E assay; **(C)** ddPCR of the diluted melanoma cancer cfDNA sample in the healthy plasma background at an expected circulating tumor allele frequency of 0.1%.

**Fig. S3.**
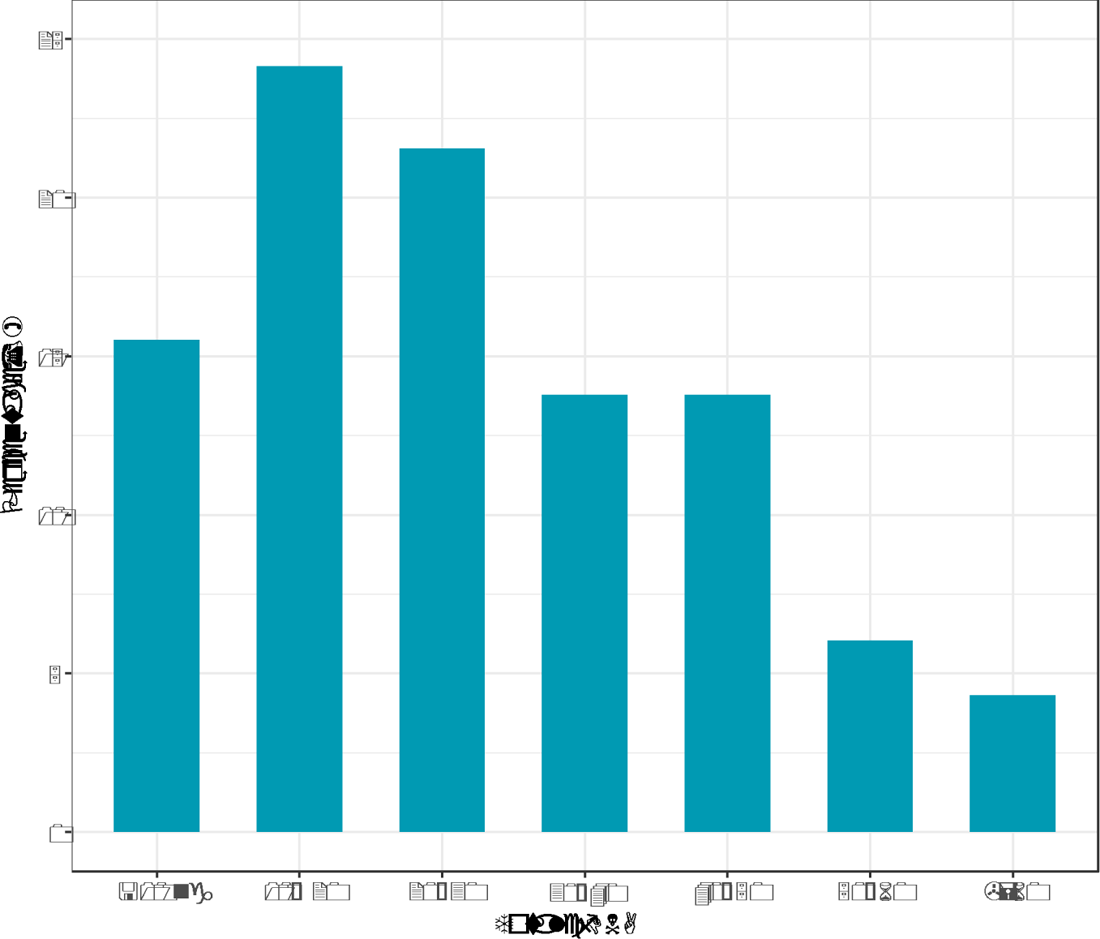
Histogram of total cell-free DNA yield from plasma samples used in this study. cfDNA was extracted from 1-4 mL of plasma.

**Fig. S4.**
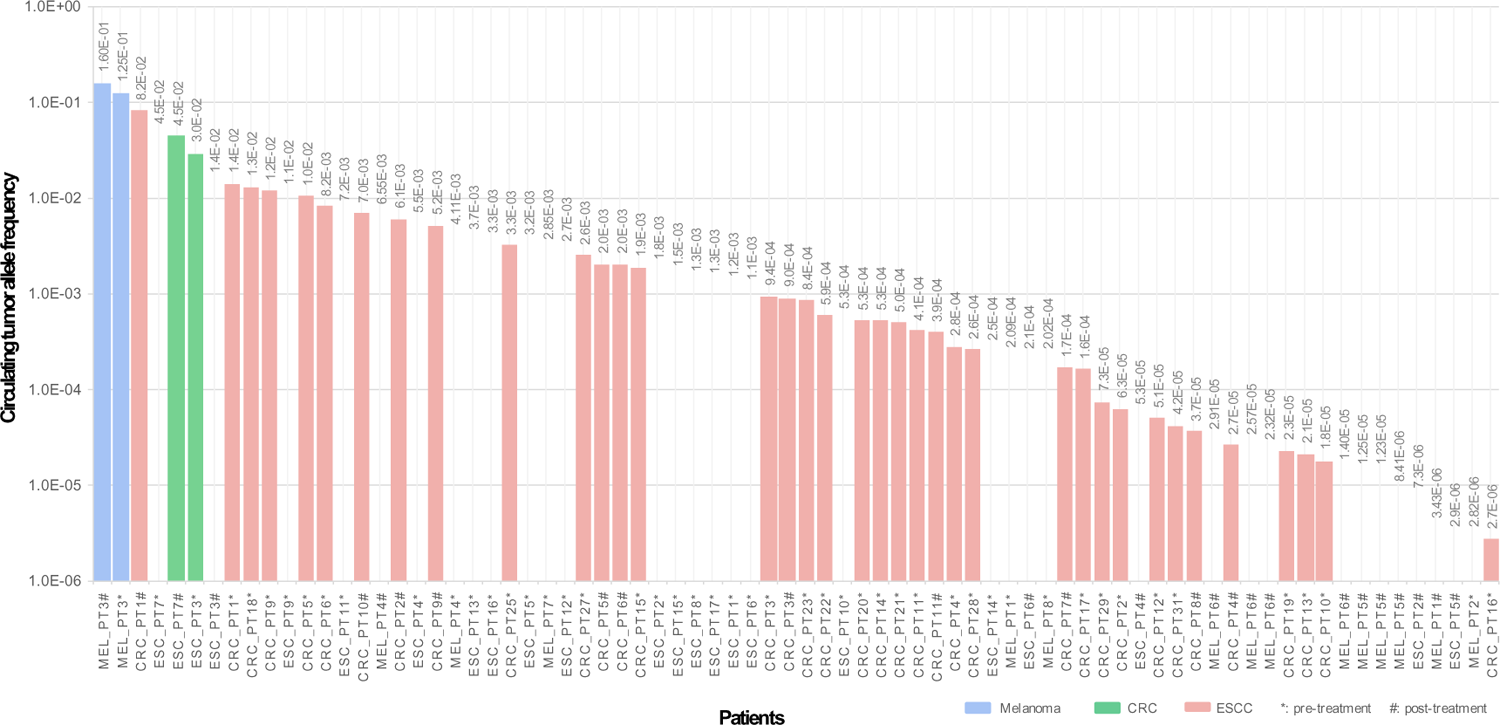
Circulating tumor allele frequency of all ctDNA positive plasma samples. Plot included pre-treatment and post-treatment samples CRC, colorectal cancer; ESCC esophageal squamous cell carcinoma.

**Fig. S5.**
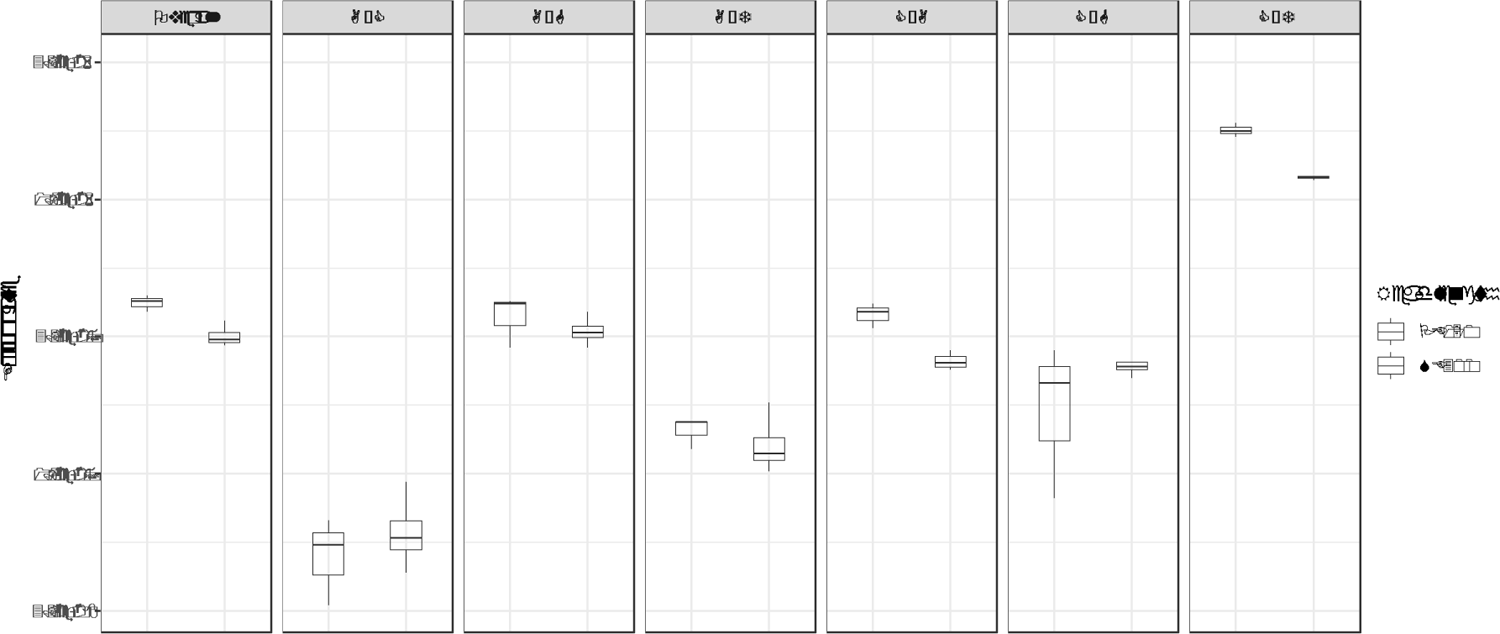
AccuScan error rate. Overall error rate and error rate of each variant type from AccuScan whole genome sequencing data on healthy human cell-free DNA samples (N=3) sequenced by pair end 150 (PE150) read length or single end 300 (SE300) read length using the 300 cycle sequencing reagents.

### Supplementary tables

**Table S1.** Different Single nucleotide polymorphic sites detected in the samples.

| **Donor** | **Recipient** | **SNP sites** |
| --- | --- | --- |
| healthy individual 1 | healthy individual 4 | 123,395 |
| healthy individual 2 | healthy individual 4 | 164,328 |
| healthy individual 3 | healthy individual 4 | 125,536 |

**Table S2.** Patient samples used in the WBC dependent and WBC-free MRD.

| Cancer type | Patient ID |
| --- | --- |
| CRC | CRC_pt9 |
| CRC | CRC_pt23 |
| CRC | CRC_pt13 |
| CRC | CRC_pt11 |
| CRC | CRC_pt19 |
| CRC | CRC_pt20 |
| ESCC | ESC_pt1 |
| ESCC | ESC_pt2 |
| ESCC | ESC_pt5 |
| ESCC | ESC_pt6 |
| ESCC | ESC_pt14 |
| ESCC | ESC_pt9 |
| Melanoma | MEL_pt7 |
| Melanoma | MEL_pt1 |
| Melanoma | MEL_pt6 |
| Melanoma | MEL_pt5 |
| Melanoma | MEL_pt8 |
| Melanoma | MEL_pt9 |
| Melanoma | MEL_pt2 |
| Melanoma | MEL_pt4 |

WBC, white blood cell; MRD, molecular residual disease; CRC, colorectal cancer;
ESCC, esophageal squamous cell carcinoma.

**Table S3.** Patients and samples information of colorectal patients.

| **Patient ID** | **Tumor tissue** | **WBC sampling** | **Pre-operative plasma sampling** | **Post-operative plasma sampling** | **Age** | **Tumor location** | **Cancer stage** | **Pre-operative ctDNA Status** | **Post-operative ctDNA Status** |
| --- | --- | --- | --- | --- | --- | --- | --- | --- | --- |
| CRC_pt1 | Y | Y | Y | Y | NA | Left | IV | Positive | Positive |
| CRC_pt2 | Y | N | Y | Y | 50-59 | Left | IV | Positive | Positive |
| CRC_pt3 | Y | N | Y | Y | <40 | Left | III | Positive | Positive |
| CRC_pt4 | Y | N | Y | Y | ≥70 | Left | III | Positive | Positive |
| CRC_pt5 | Y | N | Y | Y | 50-59 | Left | III | Positive | Positive |
| CRC_pt6 | Y | N | Y | Y | 50-59 | Left | III | Positive | Positive |
| CRC_pt7 | Y | N | N | Y | 50-59 | Left | III | Not available | Positive |
| CRC_pt8 | Y | N | N | Y | 50-59 | Left | III | Not available | Positive |
| CRC_pt9 | Y | Y | Y | Y | 50-59 | Right | I | Positive | Positive |
| CRC_pt10 | Y | N | Y | Y | 60-69 | Left | I | Positive | Positive |
| CRC_pt11 | Y | Y | Y | Y | <40 | Left | I | Positive | Positive |
| CRC_pt12 | Y | Y | Y | Y | 60-69 | Right | III | Positive | Negative |
| CRC_pt13 | Y | Y | Y | Y | 60-69 | Right | II | Positive | Negative |
| CRC_pt14 | Y | Y | Y | Y | 60-69 | Right | II | Positive | Negative |
| CRC_pt15 | Y | Y | Y | Y | ≥70 | Right | II | Positive | Negative |
| CRC_pt16 | Y | Y | Y | Y | 60-69 | Right | II | Positive | Negative |
| CRC_pt17 | Y | Y | Y | Y | 40-49 | Right | II | Positive | Negative |
| CRC_pt18 | Y | Y | Y | Y | ≥70 | Left | II | Positive | Negative |
| CRC_pt19 | Y | Y | Y | Y | 50-59 | Left | I | Positive | Negative |
| CRC_pt20 | Y | Y | Y | Y | ≥70 | Left | I | Positive | Negative |
| CRC_pt21 | Y | Y | Y | Y | 60-69 | Left | I | Positive | Negative |
| CRC_pt22 | Y | Y | Y | Y | ≥70 | Right | III | Positive | Negative |
| CRC_pt23 | Y | Y | Y | Y | ≥70 | Right | II | Positive | Negative |
| CRC_pt24 | Y | N | N | Y | 50-59 | Left | III | Not available | Negative |
| CRC_pt25 | Y | N | Y | Y | 60-69 | Left | II | Positive | Negative |
| CRC_pt26 | Y | N | N | Y | 50-59 | Left | III | Not available | Negative |
| CRC_pt27 | Y | N | Y | Y | ≥70 | Right | II | Positive | Negative |
| CRC_pt28 | Y | N | Y | Y | 60-69 | Left | II | Positive | Negative |
| CRC_pt29 | Y | N | Y | Y | 60-69 | Right | II | Positive | Negative |
| CRC_pt30 | Y | N | N | Y | <40 | Left | II | Not available | Negative |
| CRC_pt31 | Y | N | Y | Y | 50-59 | Left | I | Positive | Negative |
| CRC_pt32 | Y | N | N | Y | 50-59 | Left | III | Not available | Negative |

WBC, white blood cell; CRC, colorectal cancer; ctDNA, circulating tumor DNA; NA, not available.

**Table S4.** Patients and samples information of esophageal patients.

| Patient ID | Age | Tumor location | Cancer stage | Pre-operative ctDNA Status | Post-operative ctDNA Status |
| --- | --- | --- | --- | --- | --- |
| ESC_pt1 | 50-59 | Middle | IIIA | Positive | Negative |
| ESC_pt2 | 60-69 | Lower | IIIB | Positive | Positive |
| ESC_pt3 | ≥70 | Lower | IIB | Positive | Positive |
| ESC_pt4 | 60-69 | Middle | IIIB | Positive | Positive |
| ESC_pt5 | 60-69 | Lower | IIB | Positive | Positive |
| ESC_pt6 | 60-69 | Middle | IIB | Positive | Positive |
| ESC_pt7 | 50-59 | Lower | IIIA | Positive | Positive |
| ESC_pt8 | ≥70 | Middle | IB | Positive | Negative |
| ESC_pt9 | ≥70 | Middle | IIB | Positive | Negative |
| ESC_pt10 | ≥70 | Middle | IB | Positive | Negative |
| ESC_pt11 | 60-69 | Upper | IIIA | Positive | Negative |
| ESC_pt12 | ≥70 | Lower | IIA | Positive | Negative |
| ESC_pt14 | 50-59 | Middle | IIB | Positive | Negative |
| ESC_pt15 | 60-69 | Middle | IIB | Positive | Negative |
| ESC_pt17 | 60-69 | Lower | IIB | Positive | Negative |
| ESC_pt13 | 60-69 | Middle | IB | Positive | Negative |
| ESC_pt16 | 50-59 | Middle | IIB | Positive | Negative |

ctDNA, circulating tumor DNA.

**Table S5.** Clinical characteristics of melanoma patients.

| Patient ID | Pre-treatment<br>plasma samples | Age | Stage | Melanoma Type | Primary Location |
| --- | --- | --- | --- | --- | --- |
| MEL_pt1 | Y | ≥70 | IIIB | unknown | unknown |
| MEL_pt2 | Y | 60-69 | IIIC | sun exposed | left cutaneous nasolabial fold, desmoplastic |
| MEL_pt3 | Y | 60-69 | IV | sun exposed | left Thigh |
| MEL_pt4 | Y | 50-59 | IV | sun exposed | left Arm |
| MEL_pt5 | N | ≥70 | IIIC | sun exposed | cheek |
| MEL_pt6 | N | ≥70 | IV | sun exposed | anal skin not mucosal |
| MEL_pt7 | Y | ≥70 | IIIB | sun exposed | back |
| MEL_pt8 | Y | 40-49 | IIIC | sun exposed | back |

**Table S6.**
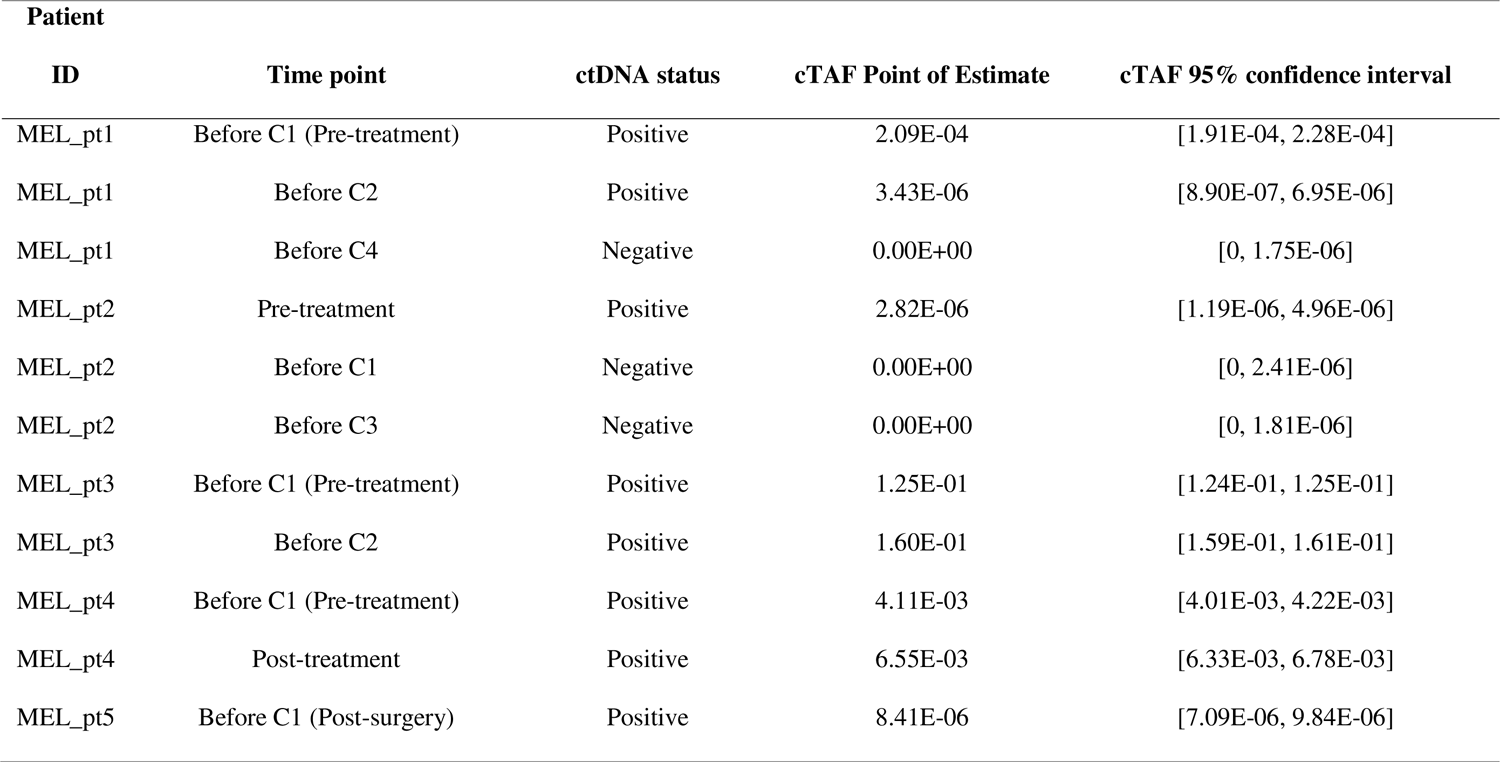
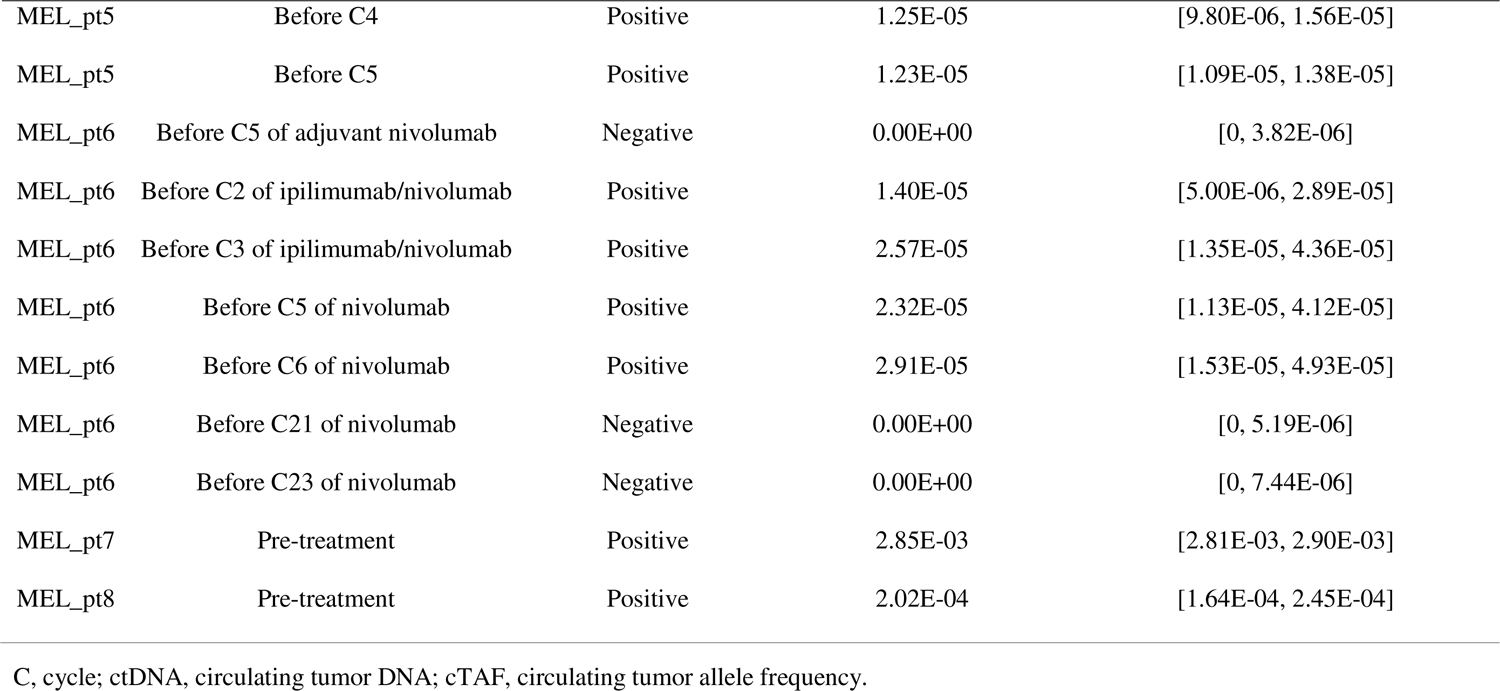
Plasma results of melanoma patients.

**Table S7.** Total cell-free DNA extracted from plasma samples.

| **Cancer type** | **Patient ID** | **Total pre-treatment cfDNA extracted (ng)** | **Total post-treatment cfDNA-1 extracted time-1 (ng)** | **Total post-cfDNA extracted time-2 (ng)** | **Total post-cfDNA extracted time-3 (ng)** | **Total post-cfDNA extracted time-4 (ng)** | **Total post-cfDNA extracted time-5 (ng)** |
| --- | --- | --- | --- | --- | --- | --- | --- |
| ESCC | ESC_pt8 | 12.56 | 36.57 | / | / | / | / |
| ESCC | ESC_pt10 | 48.24 | 25.30 | / | / | / | / |
| ESCC | ESC_pt4 | 35.04 | 46.00 | / | / | / | / |
| ESCC | ESC_pt11 | 29.28 | 28.75 | / | / | / | / |
| ESCC | ESC_pt1 | 55.20 | 24.24 | / | / | / | / |
| ESCC | ESC_pt2 | 47.52 | 42.72 | / | / | / | / |
| ESCC | ESC_pt16 | 9.11 | 27.60 | / | / | / | / |
| ESCC | ESC_pt3 | 30.72 | 52.44 | / | / | / | / |
| ESCC | ESC_pt17 | 41.76 | 43.68 | / | / | / | / |
| ESCC | ESC_pt13 | 41.52 | 42.32 | / | / | / | / |
| ESCC | ESC_pt6 | 46.32 | 50.14 | / | / | / | / |
| ESCC | ESC_pt9 | 35.04 | 37.49 | / | / | / | / |
| ESCC | ESC_pt5 | 26.68 | 51.06 | / | / | / | / |
| ESCC | ESC_pt7 | 15.59 | 23.23 | / | / | / | / |
| ESCC | ESC_pt12 | 16.74 | 33.12 | / | / | / | / |
| ESCC | ESC_pt14 | 33.36 | 60.00 | / | / | / | / |
| ESCC | ESC_pt15 | 55.20 | 30.00 | / | / | / | / |
| CRC | CRC_pt21 | 24.84 | 32.66 | / | / | / | / |
| CRC | CRC_pt9 | 46.92 | 52.08 | / | / | / | / |
| CRC | CRC_pt22 | 28.98 | 45.54 | / | / | / | / |
| CRC | CRC_pt23 | 40.02 | 59.99 | / | / | / | / |
| CRC | CRC_pt12 | 31.51 | 39.12 | / | / | / | / |
| CRC | CRC_pt13 | 15.20 | 41.86 | / | / | / | / |
| CRC | CRC_pt14 | 24.38 | 38.64 | / | / | / | / |
| CRC | CRC_pt15 | 31.68 | 24.15 | / | / | / | / |
| CRC | CRC_pt11 | 20.47 | 30.00 | 28.3 | / | / | / |
| CRC | CRC_pt16 | 37.68 | 60.00 | / | / | / | / |
| CRC | CRC_pt17 | 43.70 | 23.92 | / | / | / | / |
| CRC | CRC_pt18 | 10.00 | 34.56 | / | / | / | / |
| CRC | CRC_pt19 | 16.38 | 44.88 | / | / | / | / |
| CRC | CRC_pt20 | 48.76 | 60.00 | / | / | / | / |
| CRC | CRC_pt1 | 89.00 | 16.99 | / | / | / | / |
| CRC | CRC_pt4 | 14.54 | 10.00 | / | / | / | / |
| CRC | CRC_pt10 | 28.44 | 15.21 | / | / | / | / |
| CRC | CRC_pt8 | NA | 10.00 | / | / | / | / |
| CRC | CRC_pt5 | 9.32 | 19.54 | / | / | / | / |
| CRC | CRC_pt7 | NA | 21.21 | / | / | / | / |
| CRC | CRC_pt3 | 28.38 | 24.19 | / | / | / | / |
| CRC | CRC_pt2 | 25.02 | 17.85 | / | / | / | / |
| CRC | CRC_pt6 | 18.04 | 10.00 | / | / | / | / |
| CRC | CRC_pt25 | 12.24 | 26.64 | / | / | / | / |
| CRC | CRC_pt26 | NA | 16.33 | / | / | / | / |
| CRC | CRC_pt27 | 21.82 | 16.56 | / | / | / | / |
| CRC | CRC_pt28 | 6.86 | 5.95 | / | / | / | / |
| CRC | CRC_pt29 | 23.59 | 17.93 | / | / | / | / |
| CRC | CRC_pt30 | NA | 8.26 | / | / | / | / |
| CRC | CRC_pt31 | 6.41 | 17.65 | / | / | / | / |
| CRC | CRC_pt24 | NA | 10.00 | / | / | / | / |
| CRC | CRC_pt32 | NA | 18.15 | 7.02 | 20.01 | / | / |
| Melanoma | MEL_pt7 | 5.58 | / | / | / | / | / |
| Melanoma | MEL_pt1 | 8.42 | 8.15 | 9.00 | / | / | / |
| Melanoma | MEL_pt6 | NA | 6.53 | 10.7 | 8.00 | 12.1 | 8.30 |
| Melanoma | MEL_pt5 | NA | 9.13 | 20.7 | 15.2 | / | / |
| Melanoma | MEL_pt8 | 12.78 | / | / | / | / | / |
| Melanoma | MEL_pt2 | 5.40 | 4.82 | 6.60 | / | / | / |
| Melanoma | MEL_pt3 | 15.57 | 25.80 | / | / | / | / |
| Melanoma | MEL_pt4 | 12.89 | 66.72 | / | / | / | / |

ESCC, esophageal squamous cell carcinoma; CRC, colorectal cancer; cfDNA, cell-free DNA; NA, not available.

